# The cost of arteriovenous fistula placement in patients with chronic end-stage renal disease in Ouagadougou (Burkina Faso) 2020

**DOI:** 10.1101/2024.02.06.24302436

**Authors:** Amadou Oury Toure, Tapsoba Yanne, Fadima Yaya Bocoum, Seni Kouanda

## Abstract

End-stage renal disease (ESRD) is a late diagnosis. Its prevalence is rapidly increasing worldwide. Although various management techniques exist, they all require access routes. The arteriovenous fistula (AVF) is the vascular access of choice for regular and prolonged hemodialysis sessions. Studies have shown that an AVF saves patients money. In our context, we found no data in the literature on the cost of AVF and the lump-sum cost. The aim of this study was to determine the average costs of AVF and lump-sum costs, and to describe patients’ coping strategies in relation to these costs.

**Methods:** We conducted a cross-sectional study in the three public hemodialysis units of the city of Ouagadougou, Burkina Faso. Hemodialysis patients suffering from chronic renal failure, hospitalized or not, minors or adults having given their consent (assent) were included in the study.

**Results:** A total of 290 patients ranging in age from 12 to 82 years participated in the study. Almost half the patients (47.5%) had no income. More than half the patients had undergone fistula repair privately. Hypertension and diabetes were the pathologies most frequently associated with end-stage renal disease in this study, with 77% and 16.90% respectively. The average cost of a fistula was 260,798 Fcfa, while the average cost of a package was 506,459 Fcfa. To cope with these costs, some patients resorted to selling goods (means of subsistence) in 4.83%, borrowing, bartering and others begging.

**Conclusion:** The average cost of arteriovenous fistula and the fixed fee for haemodialysis remain an economic barrier for patients and families to haemodialysis in Burkina Faso. More than half of all patients have their fistulas done in the private sector, which costs twice as much as the public sector. Training providers working in public haemodialysis units to perform arteriovenous fistula is necessary to reduce this cost for patients.

## Introduction

End-stage renal disease (ESRD) is a late diagnosis. People at risk of CKD should be referred to a nephrology service for proper management [1]. Its prevalence is rapidly increasing worldwide [2]. It was 10.8% in China, 10.0% in the USA and 10.2% in Norway. Although various techniques are available for the management of end-stage renal failure [3-5], they require different access routes. The permanent arteriovenous fistula (AVF) is the vascular access of choice for regular and prolonged haemodialysis sessions, offering good quality, optimal flow and improved survival for haemodialysis patients [6,7]. The prevalence of AVF was over 90% in 2013 in Japan and Russia, in the USA it had increased from 24% to 68%, and in South Africa it was 51%. Studies have shown the cost of dialysis borne by patients and families [8,9]. According to the National Kidney Foundation, in the United States, Medicare spends an average of 90,000 USD per patient per year on hemodialysis treatment for CKD [10]. Another study proved that performing AVF saved beneficiaries $199 per limb per month [11]. Yang.S et al in 2017 had reported in their study an average first-year cost per patient of USD 11,240 less for AVF than for SAVF [12]. In the DRC, Izeidi.PPM et al reported in their study an average cost of catheter placement of 277.2 USD ± 134.9 USD. In addition, they found a difference between temporary and permanent catheters of US$265 and US$900 respectively [8].

In the context of Burkina Faso, we found no data in the literature on the cost of arteriovenous fistula placement and the lump-sum cost, which are prerequisites for hemodialysis in CKD patients. The aim of this study was to determine the average cost of arteriovenous fistula placement and the lump-sum cost in Ouagadougou, and to describe patients’ strategies for coping with these costs, as well as the associated pathologies.

## Methods

Study period and type: This was a cross-sectional analytical study from July 1 to August 31, 2020.

Study sites: The three hemodialysis units of the three public university hospitals of Ouagadougou were our collection sites for this study.

Study population: our study population consisted of all patients with chronic renal failure who had undergone arteriovenous fistula treatment in the hemodialysis units of the Yalgado Ouédraogo, Tingandogo and Bogodogo teaching hospitals during the study period.

Inclusion criteria: all patients with chronic end-stage renal disease who had undergone at least one arteriovenous fistula for haemodialysis and who were followed up in the three public haemodialysis units of the Ouagadougou teaching hospitals (Yalgado Ouedraogo, Tingandogo and Bogodogo) were included in this study.

### Non-inclusion criteria

Patients who had not given consent to participate in the study, patients who had undergone arteriovenous fistulation but had lost their bill, patients who had undergone fistulation but whose general condition had deteriorated and who had no major companion, patients whose data were unusable (medical records, health record) and patients with mental disorders were not included in this study.

### Study variables

These were, on the one hand, the costs of fistula placement and patient registration, and, on the other, socio-demographic variables.

The sociodemographic variables are: age, gender, education level, marital status, occupation, source of care funding, patient income level, comorbidity, household size, UHC name and residence.

### Sampling and sample size

For the purposes of the study, we decided to carry out a census of all patients who met our study criteria and who had agreed to participate in the study. This led to the construction of the table below showing the sample size collected in each hospital.

**Table 1:** Breakdown of sample by teaching hospital.

| CHU | Number | % |
| --- | --- | --- |
| **Yalgado Ouédraogo** | 67 | 23,10 |
| **Bogodogo** | 133 | 45,86 |
| **Tingandogo** | 90 | 31,03 |
| **Total** | **290** | **100,00** |

### Data collection

Data were collected using a structured questionnaire. The data collection team consisted of the principal investigator and three final-year medical students. They received two days’ training on ethical aspects and the definition of the variables to be collected. The questionnaire was then pre-tested with a sample of 10 patients in the hemodialysis unit of Bogodogo University Hospital for validation. Cost collection was based on evidence of receipts and/or invoices. Interviews were conducted in French and in the local languages of Burkina Faso (More, Foulfoulde, Dioula).

### Data analysis

Data were entered using Epidata software, then exported to Stata version 15 for analysis. Descriptive statistics were performed for socio-demographic characteristics. Pearson’s chi-square and Fisher’s exact test were used to compare categorical variables. The Kruskal-Wallis test was used for asymmetrically distributed continuous quantitative variables to compare group means and categories.

### Ethical considerations

We submitted and obtained ethical approval (N°2020-8-165) from the Burkina Faso Health Research Ethics Committee to conduct this study. To comply with ethical principles, written informed consent (assent for minors) was obtained from each participant prior to the interview. Confidentiality of data collected from participants was ensured during and after the survey. All methods were applied in accordance with current guidelines and regulations.

Informed consent was obtained from adult participants and parents/guardians of all children under 16.

## Results

A total of 290 patients were interviewed for this study. The age of the participants ranged from 12 to 82 years, with a mean age of 44.3 ±14 years. The 34-44 age group was the most common. Women accounted for 40.3%, with a sex ratio (male/female) of 1.4. The most represented socio-professional category was the unemployed (26.2%), compared with 17.24% of government employees. People with no income accounted for 47.5%. Patients with secondary education represented 42.0%.

**Table 2:** Sociodemographic and socioeconomic characteristics.

|  | Number | % |
| --- | --- | --- |
| **Age (Years)** |  |  |
| 12 - 22 | 13 | 4,5 |
| 23 - 33 | 60 | 20,7 |
| 34 -44 | 77 | 26,6 |
| 45 -55 | 73 | 25,2 |
| 56 - 66 | 49 | 17 |
| 67 and over | 18 | 6,2 |
| **Sex** |  |  |
| Male | 173 | 59,7 |
| Female | 117 | 40 |
| **Occupation** |  |  |
| Public sector | 50 | 17,2 |
| Private sector | 35 | 12,1 |
| Self employment | 37 | 12,8 |
| Students/pupils | 18 | 6,2 |
| Housewives | 53 | 18,3 |
| Retired | 21 | 7,2 |
| Unemployed | 76 | 26,2 |
| **Marital status** |  |  |
| Singles | 6 | 2,1 |
| Married | 186 | 64,1 |
| Divorced / Widowed | 61 | 21,0 |
| Concubinage | 37 | 12,8 |
| **Education level** |  |  |
| Not schooling | 50 | 17,2 |
| Primary | 46 | 15,9 |
| Secondary | 122 | 42,1 |
| University | 52 | 18 |
| Schooling in arabic language | 20 | 7 |
| **Income periodicity** |  |  |
| No income | 139 | 47,6 |
| Daily | 33 | 11,4 |
| Monthly | 102 | 35,2 |
| Trimestrial | 12 | 4,1 |
| Annual | 5 | 1,7 |
| **Level of income** |  |  |
| No income | 139 | 47,9 |
| Lower than the minimum salary<br>(34 664 Fcfa) | 32 | 11,0 |
| 34 664 to 100 000 Fcfa | 31 | 10,7 |
| 100 000 to 300 000 Fcfa | 66 | 22,8 |
| 300 000 to 500 000 Fcfa | 15 | 5,2 |
| 500 000 Fcfa and More | 7 | 2,4 |

### Analysis of comorbidity associated with chronic renal failure

Among the pathologies associated with CKD in the patients who took part in the study, arterial hypertension came in first place with 77% of cases, followed by diabetes 16.90%. The study showed that 4.48% of patients suffered from other illnesses in addition to chronic renal failure. These were: hemiplegia, hepatitis B, anemia, heart failure, tuberculosis, ulcer and pulmonary embolism.

**Table 3:** Distribution of patients according to pathologies associated with chronic renal failure.

| Associated diseases | Number | % |
| --- | --- | --- |
| Nephrotic syndrome | 1 | 0,34 |
| HTA | 226 | 77,93 |
| Diabetes | 49 | 16,90 |
| Other diseases | 13 | 4,48 |
| VIH | 1 | 0.34 |

### Distribution of patients by fistula location

It remains to be seen whether certain elements, such as the fistula and the lump-sum deposit, are borne by patients. These costs appear exorbitant. The study shows that 88% of patients have their fistula performed privately.

### Analysis of fistula costs

#### Breakdown of fistula costs by place of operation

The average cost of fistula was 277,849 Fcfa in the private sector, twice as high as in the public sector. This result was statistically significant (p= 0.000) and (IC95% [251,666 ; 304,032]).

#### Average cost of a fistula in Ouagadougou and patient coping strategies

Our results show that the average cost of a fistula in Ouagadougou was 260,798 ±12,112 Fcfa. The average lump-sum cost was 506,459 Fcfa, with a Std. Err of 5,960 and a 95% CI of [494727; 518190]. Patients’ strategies for coping with costs, apart from those strictly using public and private wages, included selling goods (means of subsistence) in 4.83%, borrowing, bartering and others begging.

**Table 4:**
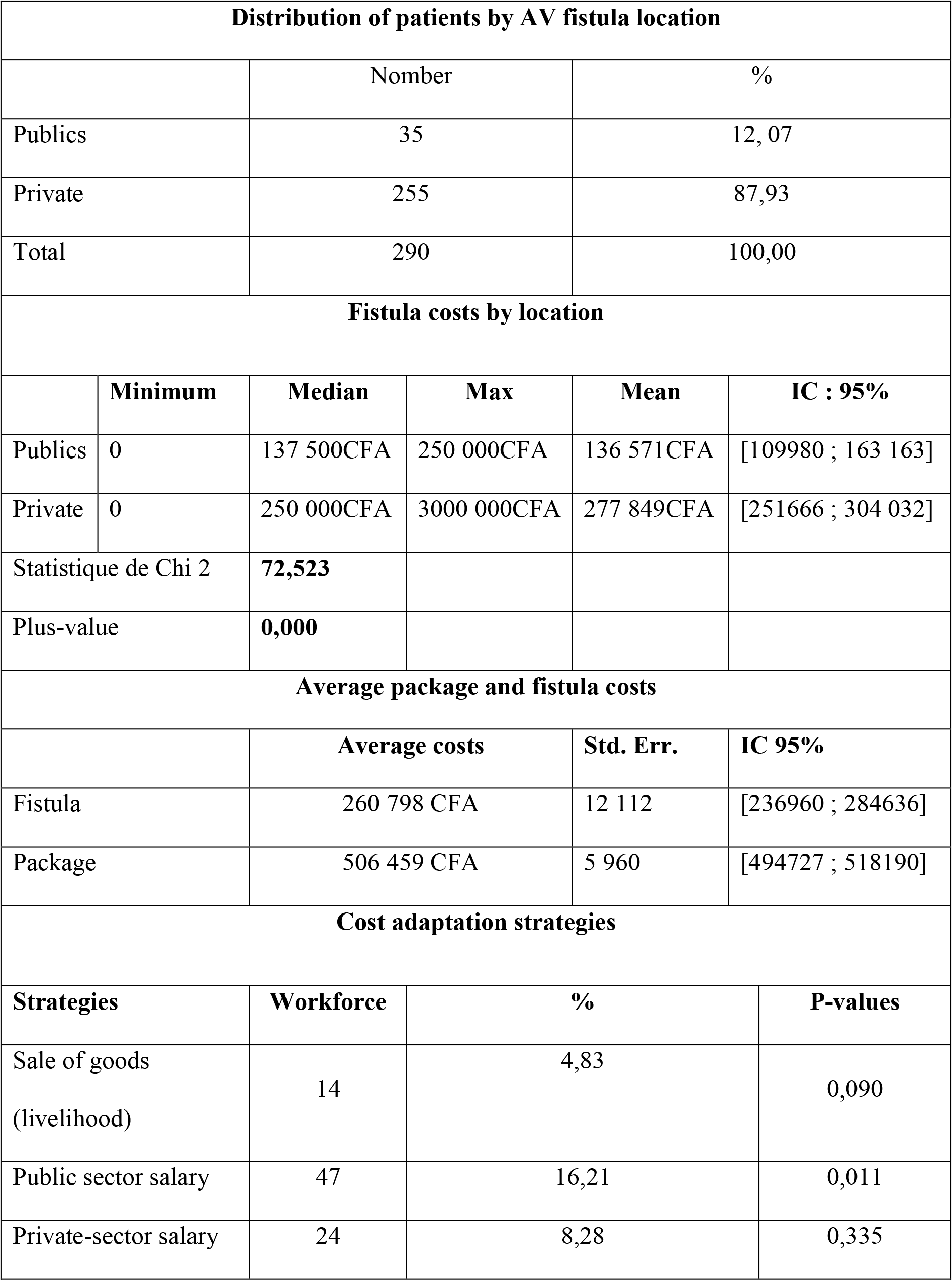
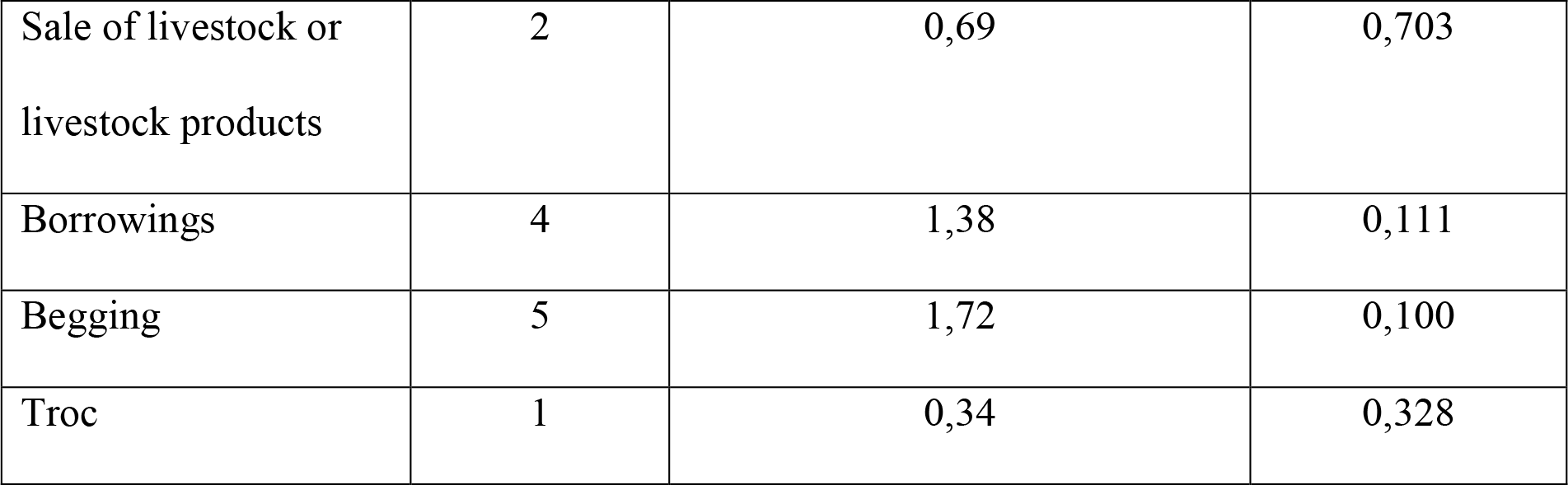
Distribution of patients according to location of AVF, cost and coping strategies.

| Distribution of patients by AV fistula location |  |  |  |  |  |
| --- | --- | --- | --- | --- | --- |
|  | Nomber |  |  | % |  |
| Publics | 35 |  |  | 12, 07 |  |
| Private | 255 |  |  | 87,93 |  |
| Total | 290 |  |  | 100,00 |  |
| Fistula costs by location |  |  |  |  |  |
|  | Minimum | Median | Max | Mean | IC : 95% |
| Publics | 0 | 137 500CFA | 250 000CFA | 136 571CFA | [109980 ; 163 163] |
| Private | 0 | 250 000CFA | 3000 000CFA | 277 849CFA | [251666 ; 304 032] |
| Statistique de Chi 2 |  | 72,523 |  |  |  |
| Plus-value |  | 0,000 |  |  |  |
| Average package and fistula costs |  |  |  |  |  |
|  |  | Average costs |  | Std. Err. | IC 95% |
| Fistula |  | 260 798 CFA |  | 12 112 | [236960 ; 284636] |
| Package |  | 506 459 CFA |  | 5 960 | [494727 ; 518190] |
| Cost adaptation strategies |  |  |  |  |  |
| Strategies |  | Workforce | % |  | P-values |
| Sale of goods<br>(livelihood) |  | 14 | 4,83 |  | 0,090 |
| Public sector salary |  | 47 | 16,21 |  | 0,011 |
| Private-sector salary |  | 24 | 8,28 |  | 0,335 |

|  |  |  |  |
| --- | --- | --- | --- |
| Sale of livestock or livestock products | 2 | 0,69 | 0,703 |
| Borrowings | 4 | 1,38 | 0,111 |
| Begging | 5 | 1,72 | 0,100 |
| Troc | 1 | 0,34 | 0,328 |

| Distribution of patients by AV fistula location |  |  |  |  |  |
| --- | --- | --- | --- | --- | --- |
|  |  | Number |  | % |  |
| Publics |  | 35 |  | 12, 07 |  |
| Private |  | 255 |  | 87,93 |  |
| Total |  | 290 |  | 100,00 |  |
| Fistula costs by location |  |  |  |  |  |
|  | Minimum | Median | Max | Mean | IC : 95% |
| Publics | 0 | 137 500CFA | 250 000CFA | 136 571CFA | [109980 ; 163 163] |
| Private | 0 | 250 000CFA | 3000 000CFA | 277 849CFA | [251666 ; 304 032] |
| Statistique de Chi 2 |  | **72,523** |  |  |  |
| Plus-value |  | **0,000** |  |  |  |
| Average package and fistula costs |  |  |  |  |  |
|  |  | Average costs | Std. Err. | IC 95% |  |
| Fistula |  | 260 798 CFA | 12 112 | [236960 ; 284636] |  |
| Package |  | 506 459 CFA | 5 960 | [494727 ; 518190] |  |
| Cost adaptation strategies |  |  |  |  |  |
| Strategies |  | Workforce | % |  | P-values |
| Sale of goods<br>(livelihood) |  | 14 | 4,83 |  | 0,090 |
| Public sector salary |  | 47 | 16,21 |  | 0,011 |
| Private-sector salary |  | 24 | 8,28 |  | 0,335 |
| Sale of livestock or<br>livestock products |  | 2 | 0,69 |  | 0,703 |
| Borrowings |  | 4 | 1,38 |  | 0,111 |
| Begging |  | 5 | 1,72 |  | 0,100 |
| Troc |  | 1 | 0,34 |  | 0,328 |

## Discussion

This study on the cost of AVF placement shows that CKD patients face an economic barrier. The cost of this activity varies according to where it is performed. The mean age of our participants was 44.3 ±14 years, ranging from 12 to 82 years. The young age in our study could be explained by the sedentary lifestyle, lifestyle and self-medication, which are factors that can influence the kidneys. A.Adedjouma et al in. 2014 had found in their study a median age of 58 years[7] . Another study carried out in Morocco by S.Cheikh et al in 2022 on the survival of the first arteriovenous fistula in chronic hemodialysis had reported a mean patient age of 46.12 ± 22 years [6]. R.R.Yemi et al in 2018 in Nigeria had found that the mean age of participants in their study was 46.3 ± 17.2 years [13].

### Analysis of comorbidity associated with chronic renal failure

Pathologies such as arterial hypertension and diabetes were the ones most associated with chronic renal failure in our study. This confirms the data in the literature.

A.Adedjouma.et al in 2014 who had found that the cause of CKD was diabetic nephropathy in 16.6%, vascular in 22.9%, glomerular in 29% and tubulointerstitial in 20.8% [7].

### Distribution of patients by fistula location

#### Analysis of fistula costs

##### Distribution of fistula costs according to location of implantation

The cost of the fistula and the deposit are also borne by patients and their families. In this study, we found an average fistula cost of 277,849 Fcfa in the private sector, which doubled the cost in the public sector (136,571 Fcfa). This result was statistically significant at p= 0.000. This may be explained by the lack of equipment in the public sector and the scarcity of specialists in this field in these facilities, which results in long patient scheduling.

In a study carried out in the Democratic Republic of Congo, Izeidi.PPM et al found the cost of vascular access to be USD 13,860 [8].

### Average cost of a fistula in Ouagadougou

#### Average package cost

To provide hemodialysis for CKD patients in Ouagadougou, patients and their families have to pay an average of 506,459 Fcfa as a lump sum. To meet these costs, patients and their families resort to selling goods (means of subsistence), borrowing, bartering and even begging. This can be explained by the high rate of unemployment and poverty in this category.

#### Study strengths and limitations

This study is the first to examine the cost of AVF and the fixed fee borne by patients in Burkina Faso. It is also one of the few studies to have analyzed patients’ coping strategies in the face of these costs. As a limitation, this study was carried out in only three hemodialysis units, whereas there are other units in the regions and in the private sector that were not included in this study.

#### Implications for research and practice

The results of this study contribute to a better understanding of the cost of AVF in public and private settings, as well as patient coping strategies for achieving hemodialysis. The results of this study will help guide actions to improve access to hemodialysis services in Burkina Faso.

They will also enable the planning of other, larger studies that will provide more important data for an understanding of the economic barriers to access to hemodialysis in Burkina Faso. Practical training in arteriovenous fistula procedures for healthcare personnel working in public hemodialysis units would be necessary to alleviate this cost and reduce waiting times.

## Conclusion

The average cost of arteriovenous fistula and the average fixed cost of hemodialysis remain an economic barrier for patients and families in the management of CKD in Burkina Faso. Despite the high cost of private fistulas, which are twice as expensive as public fistulas, and the state subsidy for public fistulas, more than half of all patients have their fistulas done privately. To reduce this cost for patients, it would be important to provide practical training for healthcare staff working in public hemodialysis units on how to perform arteriovenous fistulas, in order to shorten waiting times

## Data Availability

Data supporting the results of this study are available from [Amadou Oury Toure], but restrictions apply to the availability of these data and are therefore not publicly accessible, as our research group is working on further analyses using the same data which will then be submitted for publication. However, these data are available on reasonable request from the corresponding author [Amadou Oury Touré].

## Authors’ contributions

Study design: AOT, YT, FYB.

Data collection: AOT.

Data analysis: AOT, YT

Initial drafting of manuscript: AOT, YT and FYB

Manuscript revision: FYB, SK.

The authors have read and approved the final manuscript.

## Declarations

### Ethical approval and consent to participation

As with any study involving human subjects, this study was submitted to and approved by the Burkina Faso Health Research Ethics Committee (CERS) (N°2020-8-165). Participants were informed about the study and gave their informed consent before answering any questions.

### Consent for publication

**Not applicable**.

